# A Study of Patient-Reported Dry Eye Disease in Diabetes Mellitus in Indonesia

**DOI:** 10.1101/2024.12.05.24318300

**Authors:** Md Ikhsan Mokoagow, Nina Asrini Noor, Tri Rahayu, Damara Andalia, Niluh Archi Sri Ramandari, Marina Epriliawati

## Abstract

**Background:** Diabetes mellitus (DM), one of the most common metabolic diseases in the world, is a chronic metabolic disorder characterized by persistent hyperglycemia. In diabetic patients, ocular symptoms can develop. One commonly found ocular disease is dry eye disease (DED). DED is a multifactorial disease characterized by instability of the tear layer that causes various symptoms and/or visual impairment, usually accompanied by damage to the ocular surface.

**Objective:** This study aims to determine the prevalence of dry eye disease (DED) in type 2 DM patients and to assess the correlation between DED and various type 2 DM-related conditions in Indonesia.

**Methodology:** This study is a cross-sectional observational study. The study population includes all consecutive patients diagnosed with type 2 DM between Feb 5 and Mar 26, 2024, at three hospitals and two clinics in Jakarta. The DED assessment was conducted using the DEQ-5 questionnaire, translated and validated in Bahasa, Indonesia. The Kolmogorov-Smirnov normality test and Spearman correlation test were performed on all data sets to determine the correlation between DEQ-5 outcome and the patient’s age, disease duration, fasting blood glucose (FBG) levels, and HbA1c levels.

**Result:** A total of 309 patients were recruited in this study. The patients’ age varies from 23 to 84 years old, and the majority of them have had type 2 DM for five years or less. From 309 patients, 264 FBG and 72 HbA1c test results were recorded. Through the DEQ-5 questionnaire, 64 (20.71%) scored higher than 6, indicating they are experiencing DED. Among the factors considered, including age, gender, duration of disease, FBG level, and HbA1c level, only the FBG level correlated with DED (r_s_ = .186, n = 263, p < .002).

**Conclusion:** There’s a positive correlation between the FBG level and DED. Elevating FBG levels are associated with an increased likelihood of developing DED.

## INTRODUCTION

According to ADES (2017), dry eye disease (DED) is a multifactorial disease characterized by instability of the tear layer that causes various symptoms and/or visual impairment, usually accompanied by damage to the ocular surface.^1^ According to TFOS DEWS II (2017), DED is an eye disease caused by many factors (multifactorial) involving the eye surface, with characteristics of damage homeostasis of the lacrimal layer accompanying ocular symptoms resulting from instabilities of the eye layer, hyperosmolarity, damage and inflammation of the optical surface, as well as neurosensory abnormalities.^2–3^

Some studies in Asia show a high prevalence rate of DED of 20 - 52.4%.^1–2^ One study conducted in Riau Province, Indonesia, in 2001 showed a prevalence of 27.5%. Epidemiology, according to TFOS DEWS II, the incidence of the DED increased by about 5 to 30% in the age group over 50 years.^3^ But so far, no study in Indonesia has been done on the prevalence of national DED.

Risk factors for DED include age, race, or ethnicity; higher incidence occurs in patients of Chinese, Hispanic, Asian, and Pacific Isles descent, as well as female sex. Women are twice as susceptible to dry eye symptoms, especially those who receive estrogen hormone replacement therapy. The prevalence of dry eyes is higher in the presence of eye conditions such as blepharitis, Meibomian gland dysfunction, and conjunctive disease; in systemic conditions including arthritis, osteoporosis, urolithiasis, and thyroid disorders; and after corneal, retinal, or eye tumor surgery.^4^

Diabetes mellitus (DM) is a chronic metabolic disorder characterized by persistent hyperglycemia.^5^ It is one of the most common metabolic diseases in the world. As of 2021, there will be 537 million people with diabetes worldwide. This number is expected to increase to 643 million people in 2030, and 783 people in 2045.^6^ Type 2 DM is due to impaired insulin secretion, resistance to peripheral actions of insulin, or both. Together with other metabolic abnormalities in individuals with diabetes mellitus, persistent hyperglycemia can harm multiple organ systems, resulting in the development of debilitating and potentially fatal health complications. Microvascular complications, such as nephropathy, retinopathy, and neuropathy, are the most common, while macrovascular complications raise the risk of cardiovascular diseases by two to four times.^5^

The dry eye questionnaire DEQ-5 is a simplified version of the original DEQ.^6^ The DEQ-5 consists of five questions assessing the frequency of watery eye, discomfort, and dryness (scored on a 0–4 scale) and late-day discomfort and dryness intensity (scored on a 0 – 5 scale). DEQ-5 can discriminate between dry eye and non-dry eye patients, Sjogren’s syndrome dry eye and non-Sjogren’s syndrome dry eye, and groups with varying dry eye severity.^7^

This study has two main purposes. The primary objective is to determine the prevalence of DED among individuals with type 2 DM in Indonesia. The second objective is to assess the relationship between various type 2 DM-related factors and DED. These factors comprise the patient’s age, gender, duration of disease, most recent fasting blood glucose (FBG), and HbA1c levels, together with the outcomes of the DEQ-5 questionnaire.

## METHODOLOGY

This cross-sectional observational study gathered data through simple interviews and electronic medical records (EMR) of diabetic patients. All consecutive patients diagnosed with type 2 DM aged 18 and above are included as study subjects. The study population is taken from Feb 5 to Mar 26, 2024, at three hospitals and two clinics spread across Jakarta. The locations are Fatmawati General Hospital, JEC @ Kedoya Eye Hospital, JEC Primasana @ Tanjung Priok Eye Hospital, JEC @ Tambora Eye Clinic, and JEC @ Cinere Eye Clinic. The interview consists of several questions, such as the type 2 DM duration, latest FBG level, previous HbA1c level for the past two months, eye symptoms and other systemic comorbidities, if any, routine medication, and the DEQ-5 questionnaire. The type 2 DM duration in our study is defined as the time from the day the patient was diagnosed to the day this assessment was taken. Data taken from the EMR are demographic information (name, date of birth, and sex), and they are used to cross-check the diagnosis and routine medication taken.

Data are collected once in each location.

**Figure 1.**
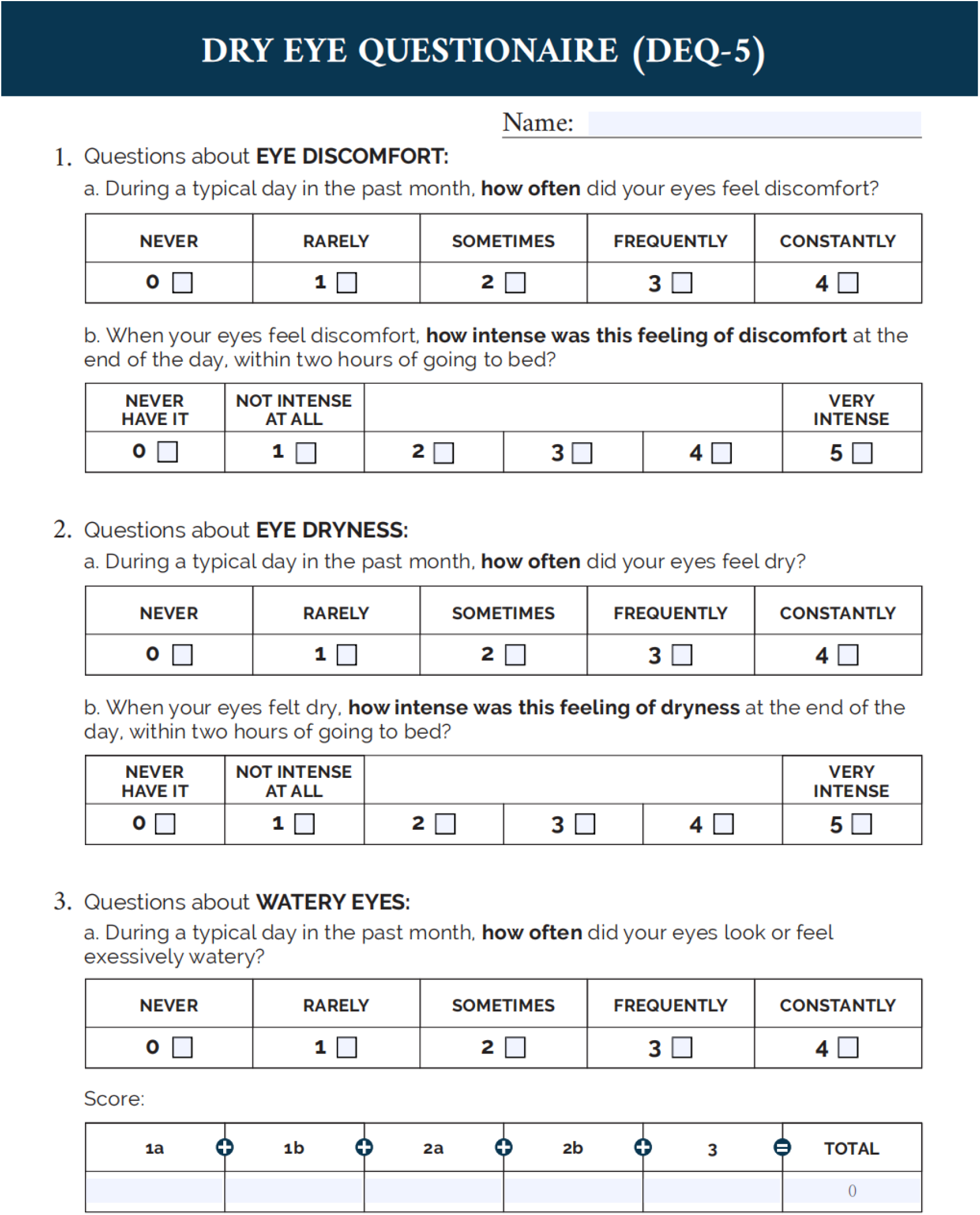
DEQ-5 dry eye questionnaire (English version).

DED assessment was conducted using the DEQ-5 questionnaire that has been translated and validated to Bahasa Indonesia. This questionnaire consists of 3 groups with five questions about dry eye symptoms. Each of the five answers will then be added to calculate the total score. The threshold value of the dry eye symptom screening criteria is DEQ-5 more than 6. The DED assessment score using the DEQ-5 questionnaire will be entered into the master table that has been prepared.

All statistics were calculated using SPSS software version 27.0 for Mac and Windows (SPSS, Inc., Chicago, IL, USA). All data underwent the Kolmogorov-Smirnov normality test to determine the data normality. The Spearman correlation test was conducted for all data sets to find the correlation between each variable and the DEQ-5 outcome.

## RESULT

A total of 309 patients were included in this study from Feb 5 to Mar 26, 2024. In the demographic data, 126 male and 184 female patients were identified. Patients’ ages varied from 23 to 84 years old, with the age group 51 – 60 years old (n = 110) being the highest proportion in this study. Most patients have been suffering from type 2 DM for less than and equal to 5 years (n = 126). The trend decreases as the duration lengthens. Their results are recorded if the patients took FBG and/or HbA1c tests. From 309 patients, 264 FBG test results are recorded. The result varies from 61 – 500 mg/dL, with most having more than 125 mg/dL (n = 146). Unfortunately, only 72 patients underwent the HbA1c test in the past two months. The results vary, with a minimum of 5.3% and a maximum of 13.5%. Most subjects (n= 29) have a 7% to <9% level. Through the DEQ-5 questionnaire, out of 309 patients, 64 (20.71%) have a total score of higher than 6.

**Table 1.**
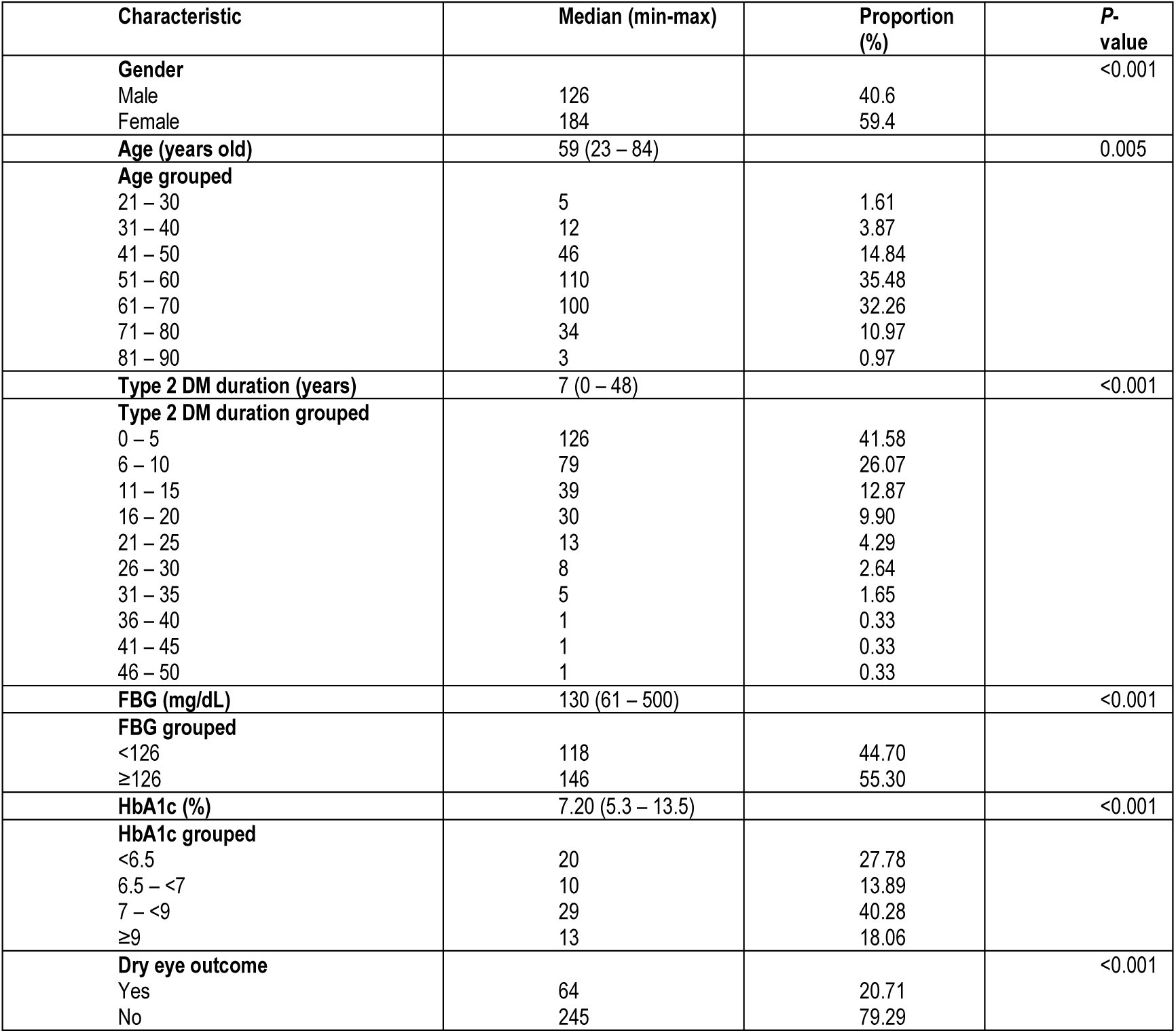
Demographics and characteristics in patients with type II diabetes mellitus (N= 309)

**Table 2.** Correlation between age group and the DEQ-5 questionnaire outcome.

| Correlations |  |  | age group<br>per 10 | DryEyeOut<br>come |
| --- | --- | --- | --- | --- |
| Spearman's<br>rho | age group per<br>10 | Correlation Coefficient | 1.000 | .009 |
|  |  | Sig. (2-tailed) | . | .879 |
|  |  | N | 309 | 309 |
|  | DryEyeOutcome | Correlation Coefficient | .009 | 1.000 |
|  |  | Sig. (2-tailed) | .879 | . |
|  |  | N | 309 | 309 |

A Spearman’s correlation was run to determine the relationship between the age group and the DEQ-5 questionnaire outcome. There was no statistically significant correlation between HbA1c level and dry eye outcome (r_s_ = .009, n = 309, p < .879).

**Table 3.**
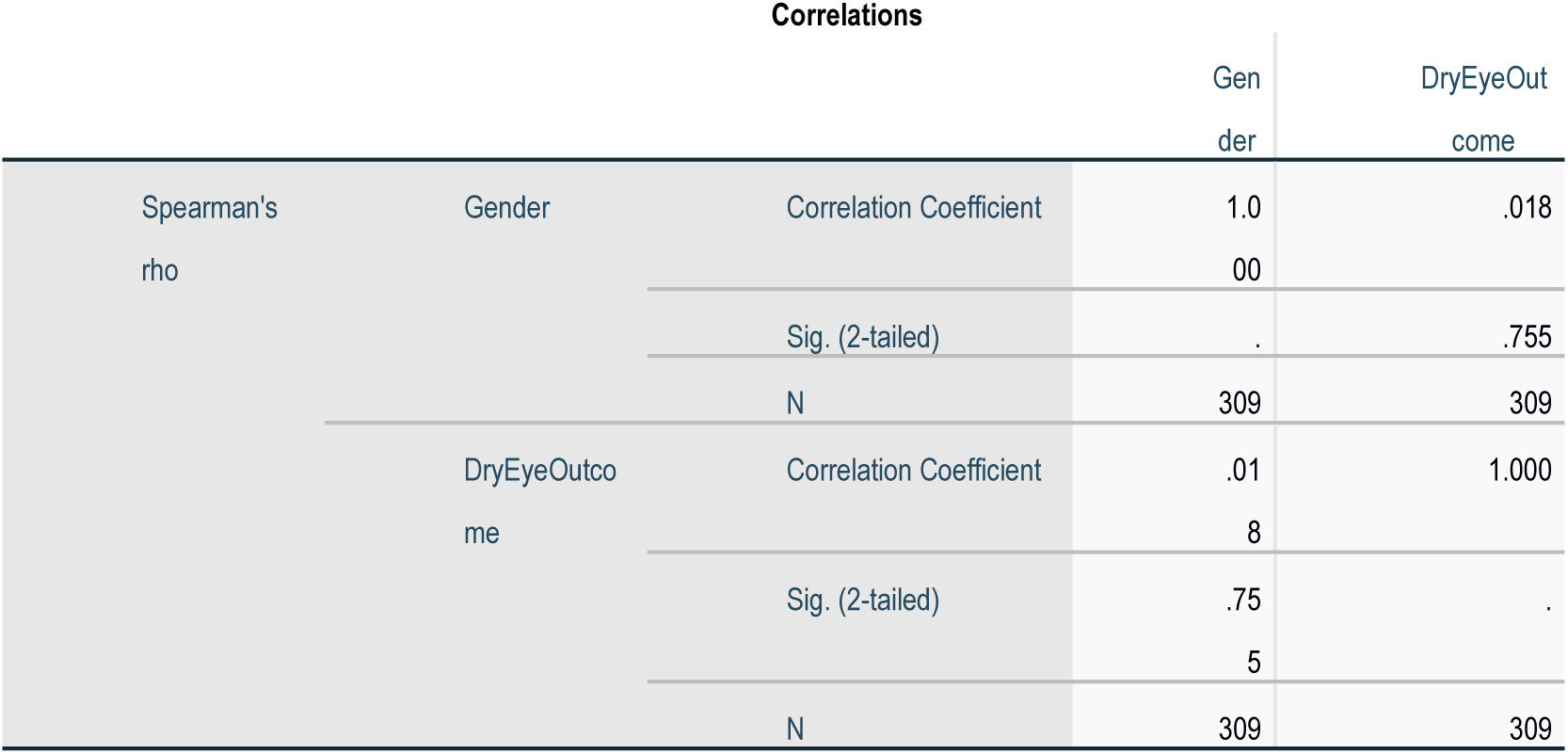
Correlation between gender and the DEQ-5 questionnaire outcome.

A Spearman’s correlation was run to determine the relationship between gender and the outcome of the DEQ-5 questionnaire. There was no statistically significant correlation between HbA1c level and dry eye outcome (r_s_ = .018, n = 309, p < .755).

**Table 4.** Correlation between duration of DM type 2 and the DEQ-5 questionnaire outcome.

| Correlations |  |  | Duration<br>per 5 years | DryEyeOut<br>come |
| --- | --- | --- | --- | --- |
| Spearman's<br>rho | Duration per 5 years | Correlation Coefficient | 1.000 | .081 |
|  |  | Sig. (2-tailed) | . | .158 |
|  |  | N | 302 | 302 |
|  | DryEyeOutcome | Correlation Coefficient | .081 | 1.000 |
|  |  | Sig. (2-tailed) | .158 | . |
|  |  | N | 302 | 309 |

A Spearman’s correlation was run to determine the relationship between the duration of suffering from type 2 DM since diagnosis and the DEQ-5 questionnaire outcome. There was no statistically significant correlation between HbA1c level and dry eye outcome (r_s_ = .081, n = 302, p < .158).

**Table 5.** Correlation between FBG level and the DEQ-5 questionnaire outcome.

| Correlations |  |  | Recent<br>FBG | DryEyeOut<br>come |
| --- | --- | --- | --- | --- |
| Spearman's<br>rho | RecentFBG | Correlation Coefficient | 1.000 | .186** |
|  |  | Sig. (2-tailed) | . | .002 |
|  |  | N | 263 | 263 |
|  | DryEyeOutco<br>me | Correlation Coefficient | .186** | 1.000 |
|  |  | Sig. (2-tailed) | .002 | . |
|  |  | N | 263 | 309 |

\*\* . Correlation is significant at the 0.01 level (2-tailed).

A Spearman’s correlation was run to determine the relationship between the FBG level and DEQ-5 questionnaire outcome. There was a weak, positive monotonic statistically significant correlation between FBG level and dry eye outcome (r_s_ = .186, n = 263, p < .002).

**Table 6.** Correlation between HbA1c level and the DEQ-5 questionnaire outcome.

| Correlations |  |  | RecentHb<br>A1c | DryEyeOut<br>come |
| --- | --- | --- | --- | --- |
| Spearman's<br>rho | RecentHbA1c | Correlation Coefficient | 1.000 | .159 |
|  |  | Sig. (2-tailed) | . | .182 |
|  |  | N | 72 | 72 |
|  | DryEyeOutco<br>me | Correlation Coefficient | .159 | 1.000 |
|  |  | Sig. (2-tailed) | .182 | . |
|  |  | N | 72 | 309 |

A Spearman’s correlation was run to determine the relationship between the HbA1c level and the DEQ-5 questionnaire outcome. There was no statistically significant correlation between HbA1c level and dry eye outcome (r_s_ = .159, n = 72, p < .182).

## DISCUSSION

Among 309 type 2 DM patients in 5 different centres in Jakarta, Indonesia, 64 (20.71%) of them are experiencing DED. This result differs from a study by Setyorini *et al*.^8^, in which out of 42 type 2 DM patients in Semarang, Indonesia, 19 (45.2%) were diagnosed with dry eye syndrome (DES). DES refers to symptoms associated with inadequate tear production or poor tear quality, while DED acknowledged the etiologies for these symptoms. DED is further classified into aqueous deficit, evaporative, and mixed etiologies.^9^ A population-based study conducted by Pan *et al*.^10^ in Taiwan identified 10,029 (6.58%) out of 152,520 patients with DED. This result proves the prevalence of DED in type 2 DM patients differs between countries.

Of all the factors considered, only the FBG level correlated with DED (r_s_ = .186, n = 263, p < .002). According to a study performed in another city in Indonesia, age, gender, and history of dyslipidemia are not significantly related to DED (p = .516, 1.000, and .155, respectively).^8^ Another study in Pakistan mentioned that gender, duration of type 2 DM, and random blood sugar level are not associated with dry eye symptoms (p = .580, .121, and .536, respectively).

Only increasing age was statistically associated with dry eye symptoms (p = .031).^11^ Contrary to this study, Pan *et al.*^10^ mentioned that DM duration is a protective factor for DED development. However, it might be masked due to underdiagnosis caused by peripheral neuropathy. The pathogenesis of dry eye caused by type 2 DM is mainly related to peripheral corneal neuropathy, tear film instability, ocular surface inflammation, and the apoptosis of conjunctival epithelial cells. Another mechanism involved is that with prolonged hyperglycemia, tear osmolarity increases, while conjunctival mucus secretion is significantly reduced, leading to decreased tear secretion and increased tear film instability.^11^

There are several limitations in this study. First, only a thorough interview with a DEQ-5 questionnaire is used to identify whether the patient has DED. Adding a Schirmer test might help establish a diagnosis. Second, unfortunately, only 72 patients had taken the HbA1c test in the last two months, which may reduce the accuracy of correlation test results.

## CONCLUSION

In type 2 DM patients, DED is associated with age, female sex, and poor glycaemic control. However, in this study, only FBG level is associated with DED. A higher FBG level increases the risk of developing DED.

## Data Availability

All data produced in the present work are contained in the manuscript.

## ACKNOWLEDGEMENTS

Ethics approval and informed consent

The study was conducted in accordance with the tenets of the Declaration of Helsinki. The authors declared no conflict of interest regarding this paper’s publication. The authors were accountable for all aspects of the work in ensuring that questions related to the accuracy or integrity of any part were appropriately investigated and resolved. The study has been authorized by the institutional ethics board of the authors, The Ethics Committee of Fatmawati General Hospital, with the ethical clearance number PP.08.02/D.XXI.18/175/2023. In compliance with institutional and national legal standards, participation in this study required written informed permission.

## Statement of Authorship

All authors certified fulfillment of ICMJE authorship criteria. The author(s) did not use an AI/AI-assisted technology in preparing, editing, and finalizing the manuscript.

## CRediT Author Statement

**MIM:** Conceptualization, Methodology, Validation, Formal analysis, Investigation, Resources, Writing – original draft preparation, Writing – review and editing, Supervision, Project administration, Funding acquisition; **NAN:** Conceptualization, Methodology, Software, Resources, Writing – original draft preparation, Writing – review and editing, Visualization; **TR:** Conceptualization, Methodology, Software, Validation, Resources, Writing – original draft preparation, Writing – review and editing, Supervision, Project administration; **DA:** Methodology, Formal analysis, Investigation, Resources, Data curation, Writing – original draft preparation, Visualization; **NAS:** Methodology, Formal analysis, Investigation, Resources, Data curation, Writing – original draft preparation, Visualization; **ME:** Methodology, Validation, Investigation, Resources, Data curation, Writing – review and editing, Project administration, Funding acquisition.

